# A FLASH model of radiolytic oxygen depletion and reactive oxygen species for differential tumor and normal-tissue response

**DOI:** 10.1101/2023.10.20.23297337

**Authors:** Jiangjun Ma, Hao Gao, Xing Shen, Xuemin Bai, Min Tang

**Affiliations:** Institute of Natural Sciences and School of Mathematics, Shanghai Jiao Tong University, Shanghai, China; Department of Radiation Oncology, University of Kansas Medical Center, Kansas City, USA; Ruijin hospital proton therapy center Shanghai Jiaotong University School of Medicine; Mevion Medical Systems, Inc., Kunshan, Jiangsu, China

## Abstract

**Objective:** FLASH-RT can potentially improve the sparing of normal tissues while preserving the tumoricidal efficiency, owing to the radiation with ultra-high dose rate. However, the FLASH mechanism remains to be solved. A popular FLASH model is based on radiolytic oxygen depletion (ROD), which explains for radiation protection of normal tissues under FLASH-RT. However, ROD does not explain the preservation of tumoricidal efficiency for tumors. This work will develop a ROS+ROD FLASH model that can explain the differential tumor and normal-tissue response.

**Approach:** The new FLASH model utilizes reactive oxygen species (ROS) in addition to ROD, and takes into account that ROS level decreases during FLASH-RT. Specifically, the differential-equation model takes into account that the basic ROS level is lower during FLASH-RT and the degeneration rates of ROS are different in tumor cells and healthy cells. Based on this ROS+ROD FLASH model, the surviving fractions of tumor and normal cells are respectively compared between conventional radiotherapy (CONV-RT) and FLASH-RT.

**Main results:** While ROD alone does not distinguish the response of tumors and normal tissues to FLASH-RT, the proposed new FLASH model based on ROD and ROS successfully explained the differential response of tumors and normal tissues to FLASH-RT, i.e., the preserved tumoricidal capability, which cannot be explained by ROD alone, and the extra normal-tissue protection owing to the ultra-high dose rate.

**Significance:** Since the ROS level decreases slower in tumors than in normal tissues, during FLASH-RT, ROS decreases more in normal tissue, thus can get more protection. By incorporating ROS in addition to ROD, the new FLASH model can not only recover all results by previous FLASH model with ROD alone, but also explain the differential response: preserved lethality of FLASH-RT to tumors and improved protection to normal tissues.

## 1. Introduction

Recent studies have suggested that, compared to conventional radiotherapy (CONV-RT) of regular dose rates, the radiation with ultra-high dose rates (FLASH-RT) can improve the normal-tissue protection while retaining the tumoricidal efficiency [1-7].

The exact mechanism behind FLASH-RT remains unclear, particularly, why FLASH-RT and CONV-RT have the same response to tumor cells hasn’t been fully investigated [36]. Pratx et al proposed that the normal-tissue sparing effect of FLASH-RT is due to the rapid reduction of intracellular oxygen during FLASH-RT, namely radiolytic oxygen depletion (ROD) [8,9]. We will refer to the FLASH model in [8] as ROD FLASH model. However, ROD does not explain why FLASH-RT is as effective as CONV-RT for killing tumor cells. Rudi Labarbe et al proposed a model to simulate the time-dependent evolution of organic peroxyl radicals *ROO*, which assumes that the reduction of *ROO* lifetime is likely to protect normal tissues during FLASH-RT [10]. In fact, according to experimental data in [33], the oxygen consumption in cells during FLASH is not as much as the setting in [8] (0.17 mm Hg/Gy rather than 1.78 mm Hg/Gy), which means that ROD alone cannot fully explain the sparing effect of FLASH-RT. So, we want to introduce ROS to explain the mechanism behind FLASH better.

This work aims to develop a new FLASH model in order to explain the differential response of tumors and normal tissues during FLASH-RT. The ROS+ROD FLASH model considers both effects of ROD and intracellular reactive oxygen species (ROS). ROS is a general term for some unstable and active oxygen-containing particles [11], including *ROO*. It plays an important role that differentiates the physiology of tumor cells from that of healthy cells [12]. The influence of ROS on the tumor development is a complex physiological process, involving multiple temporal and spatial scales [13,14], and it can be both a tumor-promoting and a tumor-suppressing agent [15].

ROS includes many different chemical ingredients. However, it is almost impossible to measure the detailed time evolution of all ingredients, thus in most mathematical tumor models, only the total density concentration [ROS] is considered instead of distinguishing between specific components in ROS. Moreover, quantitative monitoring of the time dynamical data of ROS is possible [11], which is critical to whether the ROS+ROD FLASH model shown below(model(3) and (9)) can be verified by experiments. ROS may play different roles in tumor and healthy cells. On the one hand, ROS in tumor cells are more likely to achieve oxidative stress due to the high level of basic ROS level and the lack of enzymes that can quickly degenerate ROS [16]. The increase of ROS level until it exceeds a threshold of oxidative stress will lead to cell death, which is the mechanism behind chemotherapy [17,18,19]. On the other hand, healthy cells have lower level of basic ROS level and possess enzymes that can eliminate ROS quickly [16]. Here, ‘basic ROS level’ refers to the ROS level in the cells at steady state. Therefore, the ROS degeneration rate of tumor cells is significantly slower than that of healthy cells [16].

We build a ROS+ROD FLASH model that takes ROS into account in addition to ROD, for explaining the differential response of tumor and health cells during FLASH-RT in terms of surviving fraction. The model can get the proportion of surviving cells compared to the total cell numbers at different regions after radiotherapy, that is, the space dependent surviving fraction. It can not only recover the results of the ROD FLASH model with ROD alone in [8,9], but also explain the differential response: the protection effect of FLASH-RT to normal tissue is more significant than that for tumor cells. All the results are implemented in MATLAB (version 2022a).

## 2. Methods and Materials

### 2.1. Change of oxygen during FLASH-RT

The change of oxygen during FLASH-RT is modeled via ROD [8,9]. Similar as in [8], we consider a multicellular tumor spheroid model that cells form a ball with a radius R in the center of a domain and are surrounded by a sphere shell with a radius H filled with the culture medium (Fig. 1) which is the same as in [8]. The oxygen tension at the atmospheric interface of the medium is denoted by p_air_.

**Figure 1.**
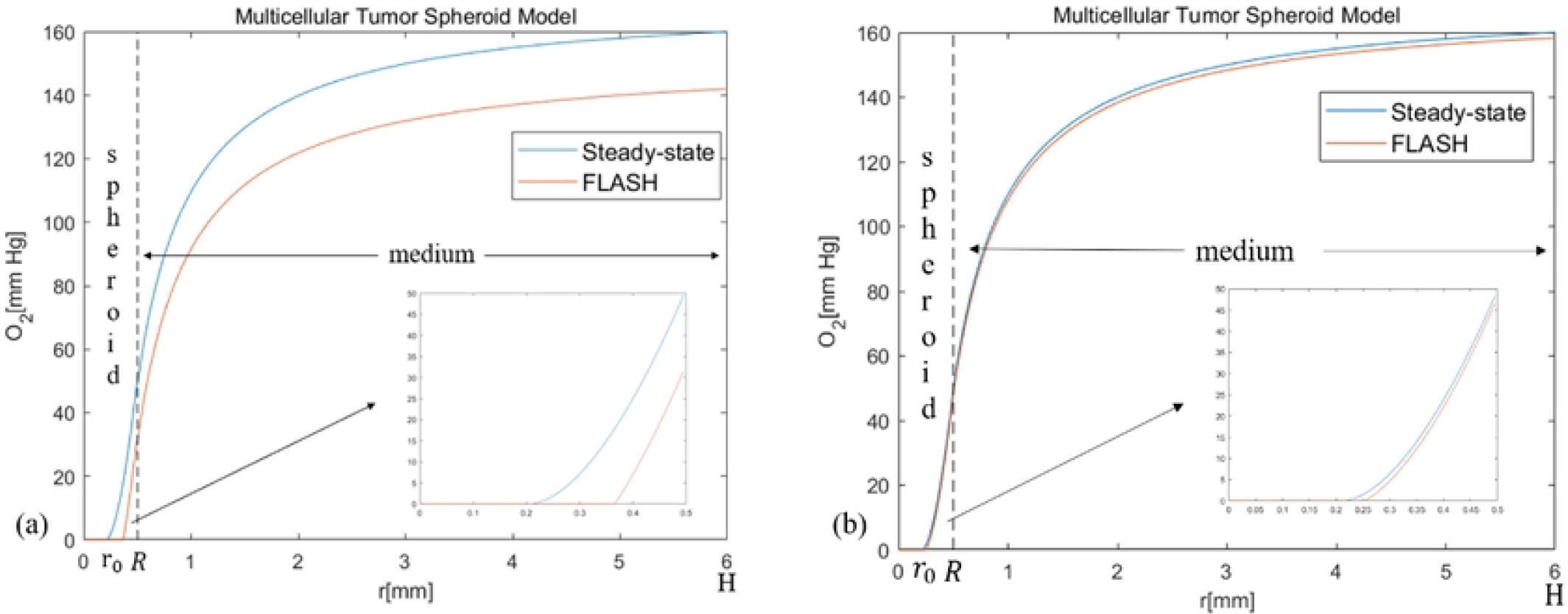
Oxygen tension inside the cell spheroid and surrounding culture medium. (a) The simulation parameters are the same as in [8]. R = 0.5 mm, H = 6 mm, 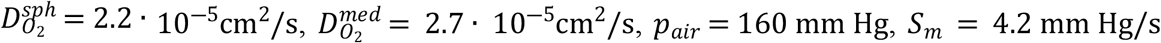 and *S*_*ROD*_ = 160mm Hg/s. (b) *S*_*ROD*_ = 15.3mm Hg/s and other parameters are the same as in (a). After FLASH-RT irradiation, the oxygen tension decreases rapidly. The original anoxic core is further expanded.

Oxygen consumption rate is set to be *S*_*m*_ inside the cell region, and zero in culture (since the culture itself does not consume oxygen). Before irradiation, the system is at a steady state and the oxygen tension is computed by solving the diffusion equation over the oxygenated spheroid and the surrounding medium [8]:

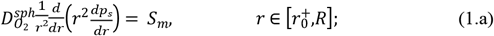

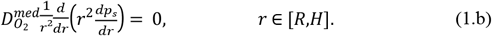

Here, *p*_*s*_ is the oxygen tension at steady state, 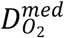 and 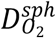 are oxygen diffusion coefficients respectively inside the medium and the cell spheroid. The oxygen concentration in the medium will change with *S*_*m*_, and the value of *S*_*m*_ here neglects the fact that tumor can be reoxygenated by vascularization. We have the continuity of *p*_*s*_(*r*) and 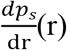 at r = R, *p*_*s*_(*H*) = *p*_*air*_ and 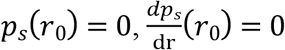. There are three boundary conditions for Eq. (1) in the interval [*r*_0_, *H*] and thus can determine not only the profile of *p*_*s*_(*r*) but also the value of *r*_0_. When 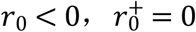 and when 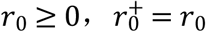. Then *p*_*s*_(*r*) ≥ 0 for 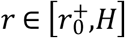 and when 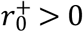, there exists an anoxic core in the cell ball (as shown in Fig. 1), which means *p*_*s*_(*r*) = 0 when *r* ∈ [0,*r*_0_]. This hypoxic region could be considered as the necrotic core where tumor cells are at quiescent state. The oxygen content of this part is very low, there is no difference between FLASH-RT and CONV-RT. However, in the proliferating region (i.e. *r* ∈ [*r*_0_,*R*]), FLASH-RT can have an obvious decline of oxygen content.

During FLASH-RT, the ultra-high-dose-rate radiation induces ROD. The oxygen tension during FLASH-RT is assumed to decrease linearly with respect to time until it reaches zero [8]. This can be modeled by:

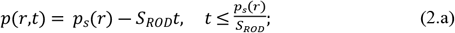

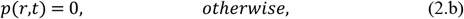

Where *p*_*s*_(*r*) is the steady-state oxygen tension in Eq. (1), *S*_*ROD*_ represents the rate of ROD. According to [8], 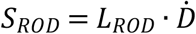 which increases with the dose rate 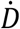, at CONV-RT dose rates, *S*_*ROD*_ can be ignored, since it its much less than the oxygen metabolism rate (*S*_*m*_ = 4.2 mm Hg/s). On the other hand, at FLASH-RT dose rates, ROD is a major contributing factor to oxygen tension. In order to fit the experimental results in [16], in [8], *S*_*ROD*_ is set to 160 mm Hg/s for FLASH-RT dose rate (90 Gy/s) and 0.02 mm Hg/s for CONV-RT dose rate (0.075 Gy/s). However, according to the experimental results in [33], in vitro experiments, during FLASH, *L*_*ROD*_ is measured to be 0.17 mm Hg/Gy, thus *S*_*ROD*_ can be measured as 15.3 mm Hg/s 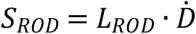. As shown in Fig.1, the oxygen tension decreases significantly after FLASH-RT irradiation, but oxygen tension decreases less when *S*_*ROD*_ becomes smaller.

### 2.2. Change of ROS during FLASH-RT

Although the change in oxygen via ROD can explain the normal-tissue protection of FLASH-RT[20,21], it does not explain the differential response to tumors, i.e., why FLASH-RT does not provide the same protection effect to tumor cells.

Our hypothesis for explaining the differential response of FLASH-RT is based on ROS. It is known that while FLASH-RT consumes oxygen rapidly, the amount of ROS produced in cells decreases as well [16], which could be another explanation of the protection effect. On the other hand, ROS in tumor cells are more likely to lead to cell death due to the high level of basic ROS level and the lack of enzymes that can quickly eliminate ROS [16].

According to [29], ROS level generated during FLASH-RT is lower than during CONV-RT. Therefore, the evolution of ROS level can be modeled by the following equations:

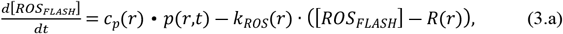

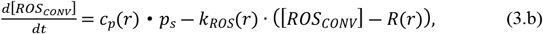

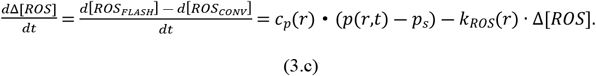

ROS can be degenerated by a series of complex biochemical reactions [10]. By [16], for simplicity, we assume that ROS is produced mainly by oxygen consumed and the degeneration rate of ROS is *k*_*ROS*_(*r*) and the equilibrium ROS level in CONV-RT is *R* (*r*), while their values may be different for tumor and healthy cells. Compared with CONV-RT, ROS level after FLASH-RT decreases. Agreement of the ROD FLASH model with the experimental data of spheroid survival fraction of [8] was achieved with a value of *S*_*ROD*_ (160mmHg/s) as calculated by Spitz et al. [16] which is much higher than the one measured in water. It is justified by incorporating lipid peroxidation and Fenton reactions inside the spheroid, which can be considered as an effect of ROS. Thus, we use simple dependence between the decreasing amount of ROS and the oxygen tension, which can be made more realistic. The equilibrium ROS level during FLASH-RT is assumed to be *R*(*r*), Since we only care about the differential response of FLASH-RT and CONV-RT to tumor cells and healthy cells, we only need to model the difference of ROS level between FLASH-RT and CONV-RT without knowing the specific value of R(r). And the ROS level difference between FLASH-RT and CONV-RT can be calculated by (3.c). Tumor cells have a higher ROS level base and a lower consumption rate than healthy cells. According to [10], *k*_*ROS*_(*r*) is set to be 0.8 · 10^―3^ *μM*^―1^ · *s*^―1^ in the tumor region and 10^―2^ *μM*^―1^ · *s*^―1^ in the normal-tissue region.

Note that for simplicity, *c*_*p*_, *k*_*ROS*_(*r*) and *R*(*r*) depend only on different cell types, while they may depend on *p*(*r,t*) and dose rate in practice, which will need to be verified through experiments regarding the ROS level during FLASH-RT.

### 2.3. FLASH model via ROD and ROS

The surviving fractions of irradiated cells are modeled by the linear-quadratic (LQ) model as in [8]. Let *N*(*r,t*) be the cell number at position r at time t. The fraction of cells 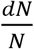 killed between time t and t + dt is

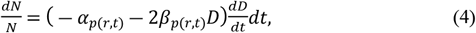

where D is the radiation dose, and 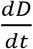 is the instantaneous dose rate. Oxygen enhancement of radiation damage occurs in a very short time which has the same time scale as FLASH-RT [23]. One needs to take the effect of oxygen tension into account. The modified parameters of the LQ model depend on the oxygen tension *p*(*r,t*) [24], i.e.,

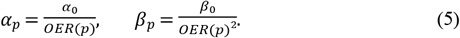

where *OER*(*p*) is the oxygen enhancement ratio (OER)[25], such that

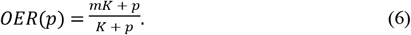

For CONV-RT, since *p*(*r,t*) ≈ *p*_*s*_(*r*), *α*_*p*(*r,t*)_ and *β*_*p*(*r,t*)_ do not change with time. Then the surviving fraction during the radiation time [0, T] is given by [8]

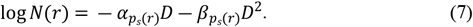

During FLASH-RT, *p*(*r,t*) changes rapidly, *α*_*p*(*r,t*)_, *β*_*p*(*r,t*)_ change with time and the surviving fraction during the radiation time [0, T] is given by [8]

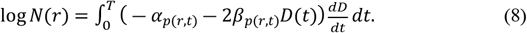

Eq. (8) gives the cell survival rate during FLASH-RT due to the decline of oxygen content, the ROS level relates to the changes of oxygen content but it depends on different cell type as well. To account for the contribution of ROS, the surviving fraction model (4) is modified as the following for the fraction of cells killed between radiation time [0,T]

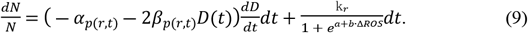

Model (9) indicates that FLASH-RT’s protective effect mainly comes from two aspects: one is radiation protection caused by decreased oxygen tension; the other is that the ROS level after FLASH-RT is lower than CONV-RT, thus reducing the oxidative stress effect of ROS.

In fact, in the latter aspect, we model the effect of Δ*ROS* on cell survival through 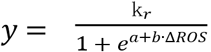, and we want y to be equal to 0 (or very close to 0) when Δ*ROS* is equal to 0. When y is a little bit away from 0, Δ*ROS* must have a significant increase, but cannot exceed a certain threshold. Only in this case can the differential response of ROS to tumor and healthy cells be reflected. A function of the shape 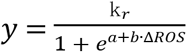, shown in Figure 2, satisfies our requirements.

**Figure 2.**
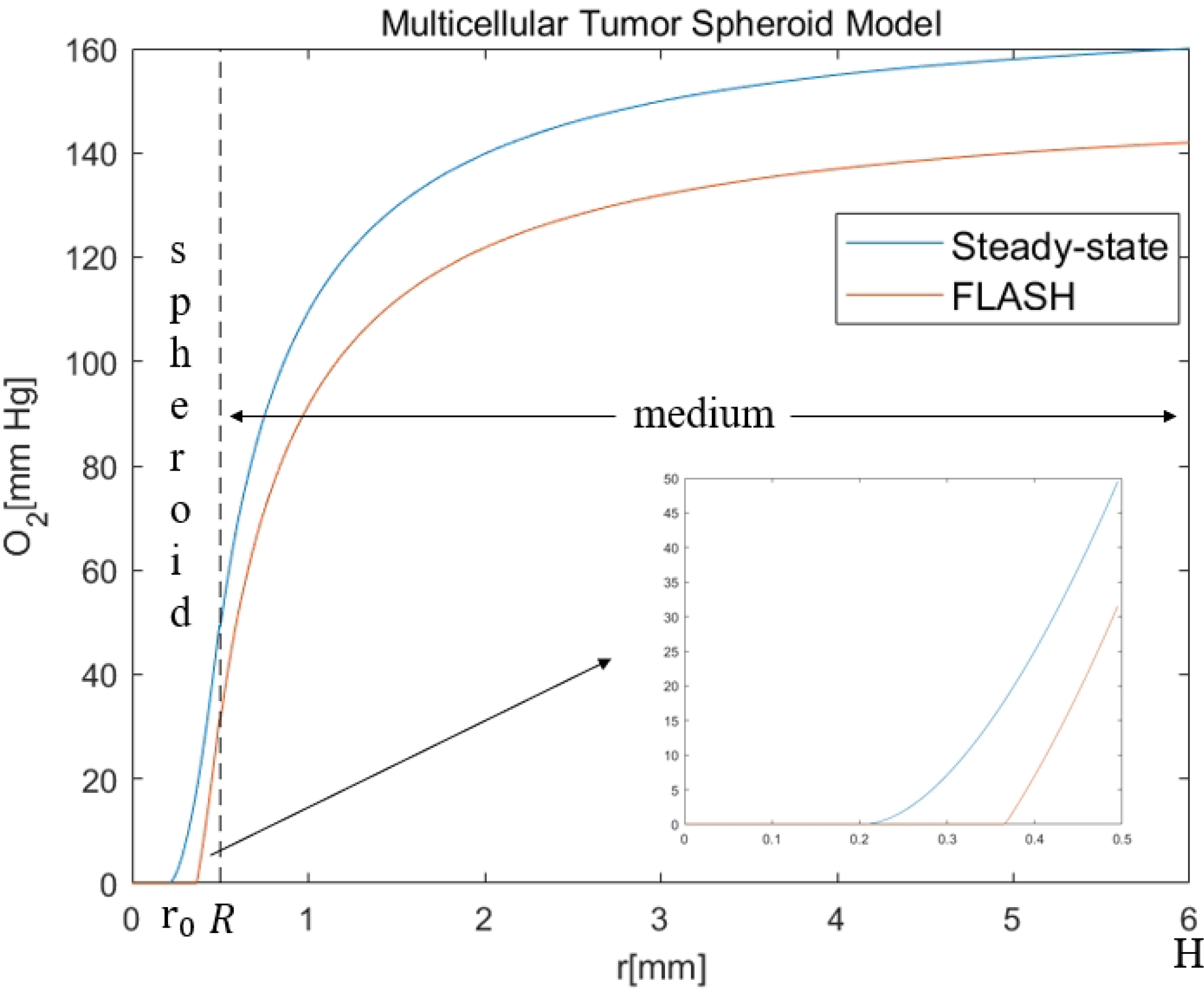
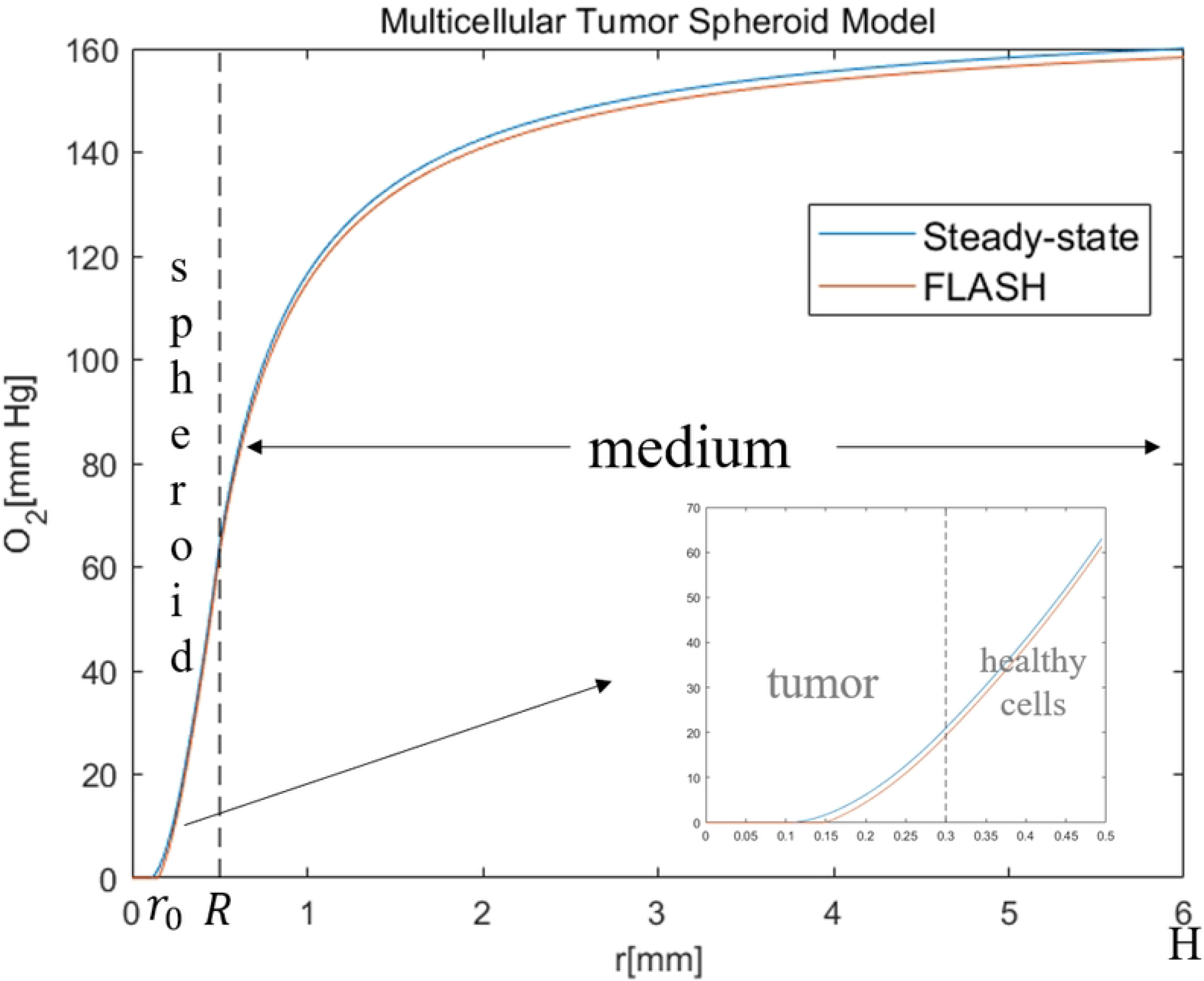
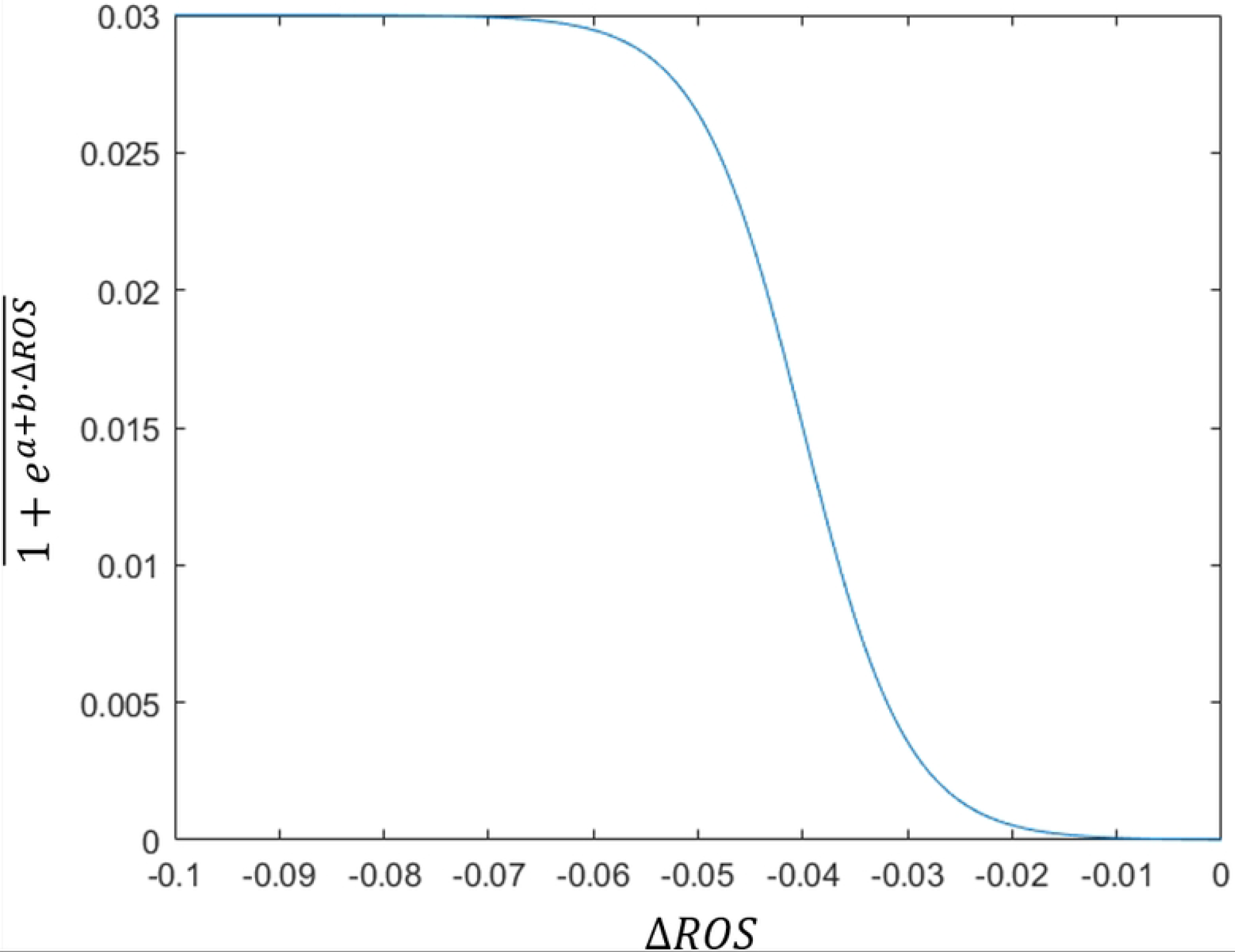
The graph of the function 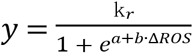 where *a* = 8, *b* = 200*μM*^―1^ and *k*_r_ = 3 · 10^―2^.

For tumor cells, due to the presence of hypoxic core, the reduction of oxygen tension after FLASH-RT is not significant. At the same time, there is not much difference between the ROS level after FLASH-RT and that after CONV-RT, so the cell surviving fraction is similar to that of CONV-RT. For healthy cells, FLASH-RT has a significant protective effect due to decreased oxygen tension and low ROS levels.

All parameters and their references for the proposed ROS+ROD FLASH model are summarized in Table 1.

**Table 1.** Parameters for the proposed ROS+ROD FLASH model via ROD and ROS.

| Notation | Parameter | Value | Reference |
| --- | --- | --- | --- |
| $D_{O_2}^{sph}$ | the rate of diffusion of oxygen in spheroid | $2.2 \cdot 10^{-5} \text{cm}^2/\text{s}$ | [27,28]<br>consistent with<br>ref [8] |
| $D_{O_2}^{med}$ | the rate of diffusion of oxygen in medium | $2.7 \cdot 10^{-5} \text{cm}^2/\text{s}$ | [26]<br>consistent with<br>ref [8] |
| $S_m$ | oxygen consumption rate | 4.2 mm Hg/s | [8]<br>consistent with<br>ref [8] |
| $S_{ROD}$ | The rate of ROD in ref [8] | 160 mm Hg/s | [16]<br>consistent with<br>ref [8] |
| $S_{ROD}$ | The rate of ROD in ROS+ROD FLASH model | 15.3 mm Hg/s | [33] * |
| $c_{p,tumor}$ | parameter in ROS model | $0.8 \cdot 10^{-4} \text{s}^{-1}$ | * |
| $c_{p,healthy}$ | parameter in ROS model | $1 \cdot 10^{-3} \text{s}^{-1}$ | * |
| $k_{ROS,tumor}$ | ROS degeneration rate in tumor cells | $0.8 \cdot 10^{-3} \text{s}^{-1}$ | [10] * |
| $k_{ROS,healthy}$ | ROS degeneration rate<br>in healthy cells | $10^{-2} s^{-1}$ | [10] |
| $m$ | OER parameter | 2.6 | [24]<br>consistent with<br>ref [8] |
| $K$ | OER parameter | 1.9 mm Hg | [24]<br>consistent with<br>ref [8] |
| $\alpha_0$ | parameter in LQ model | $0.44 Gy^{-1}$ | [8]<br>consistent with<br>ref [8] |
| $\beta_0$ | parameter in LQ model | $0.44 Gy^{-2}$ | [8]<br>consistent with<br>ref [8] |
| $k_r$ | parameter in cell<br>survival model | $3 \cdot 10^{-2}$ | * |
| $a$ | parameter in cell<br>survival model | 8 | * |
| $b$ | parameter in cell<br>survival model | $200 \mu M^{-1}$ | * |

\* The parameters were estimated by fitting the data in [8], since the exact data could not be found.

## 3. Results

### 3.1. Normal-tissue sparing of FLASH-RT via ROD only

When *S*_*ROD*_ is set to 160 mm Hg/s and *k*_*r*_=0, our model reduces to the ROD FLASH model that has been studied in [8]. First, we present the results from the ROD FLASH model and show that the results in [8] can be recovered.

Similar as in [8], we set FLASH-RT dose rate to be 90 Gy/s, CONV-RT dose rate to be 0.075 Gy/s, and the dose to be 10Gy unless noted. Moreover, we consider the multicellular tumor spheroid model with *R* = 0.5*mm* and *H* = 6*mm*. Only the results in r∈[0, R] are shown below.

Without considering ROS, the decrease of oxygen tension leads to the increase of cell surviving fraction (Fig. 3(a)), regardless of tumor or healthy cells. Considering ROD alone could explain the protective effect of FLASH-RT, but could not explain the differential response of FLASH-RT to tumor and healthy cells. Note that Fig. 3(a) was not shown in [8], but the same model and parameters are used to obtain this result. Moreover, the FLASH effect for sparing normal tissues also depended on the dose, i.e., improved cell survival as the dose increased (Fig. 3(b)), which is consistent with [8].

**Figure 3.**
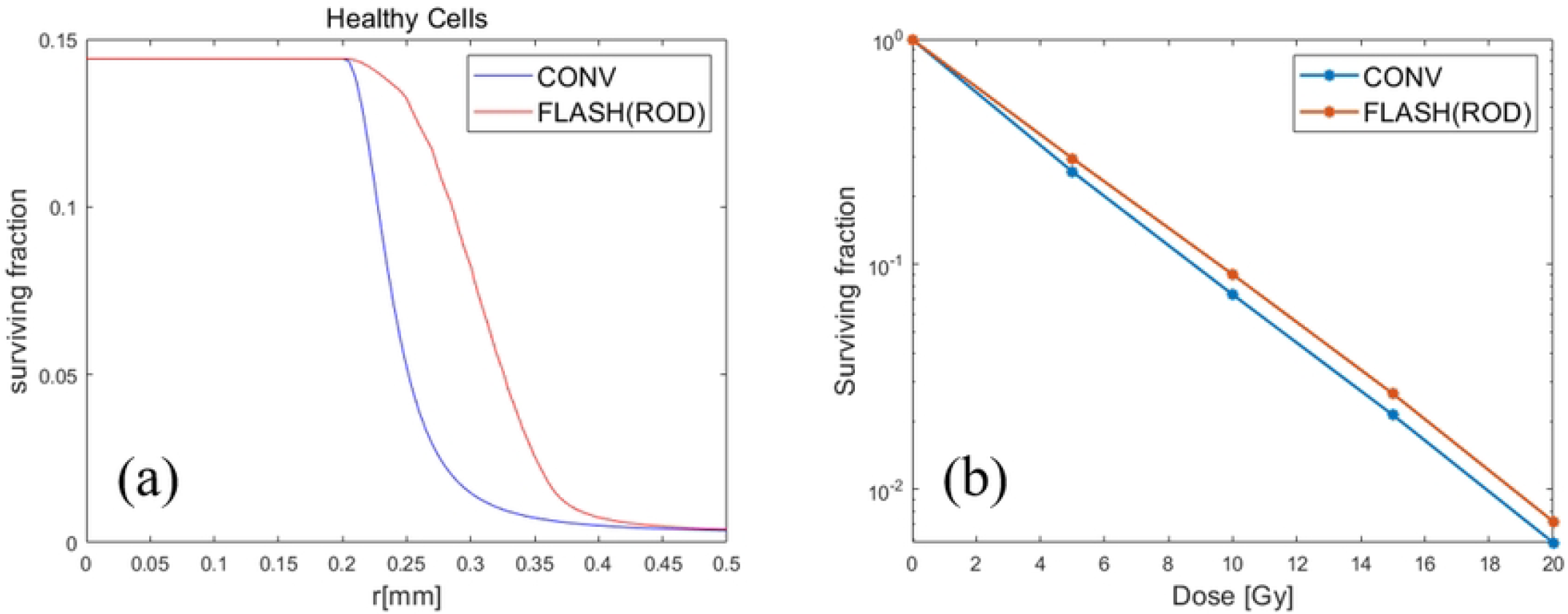
CONV-RT *v.s*. FLASH-RT with ROD only when the dose is 10Gy. (a) Compared with CONV-RT, the cell surviving fraction of FLASH-RT was significantly improved due to the radiation protection effect caused by the rapid decline of oxygen tension. However, there was no difference in surviving fraction of hypoxic core due to unchanged oxygen tension. (b) The overall cell surviving fraction (in logarithm with base 10) with respect to different dose, i.e., 5, 10, 15 and 20Gy respectively.

### 3.2. Comparison of tumor and healthy cells when ROS is considered

The same setting as in 3.1 is used but the effect of ROS is considered. In [8], the authors take *S*_*ROD*_ = 160 *mm Hg*/*s* which can be thought of as the effect of ROD plus a global ROS. Here we separate the effects of ROS on cells from ROD and consider the differential reactions of ROS in tumor and healthy cells. For the convenience of ROS calculation, the unit mm Hg of oxygen tension in Eq. (1) and (2) is converted into *μM O*_2_, i.e., from 0.77 mm Hg *O*_2_ to 1 *μM O*_2_ [26]. Moreover, according to [33], it was found in the experiment that the decrease of intracellular oxygen tension after FLASH-RT was not as much as expected in [8]. Therefore, *S*_*ROD*_ in Eq. (2) is set to be 15.3 mm Hg/s according to [16]. As in Figure 4, proper choices of the parameters *a* = 8, *b* = 200*μM*^―1^, *k*_*r*_ = 3 · 10^―2^ in (3) and (9) can reproduction the results in [8] after considering both effects of ROD and global ROS.

**Figure 4.**
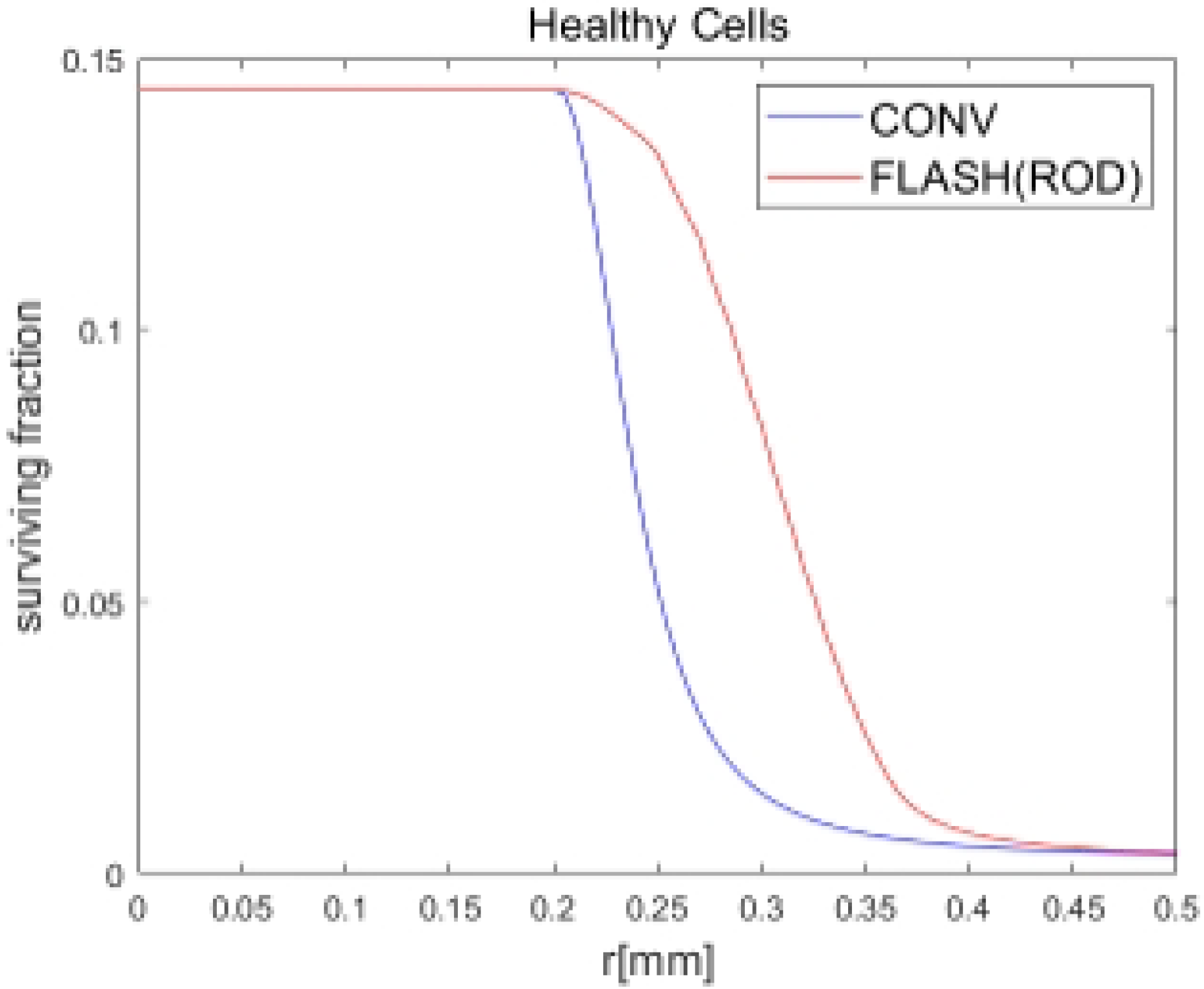
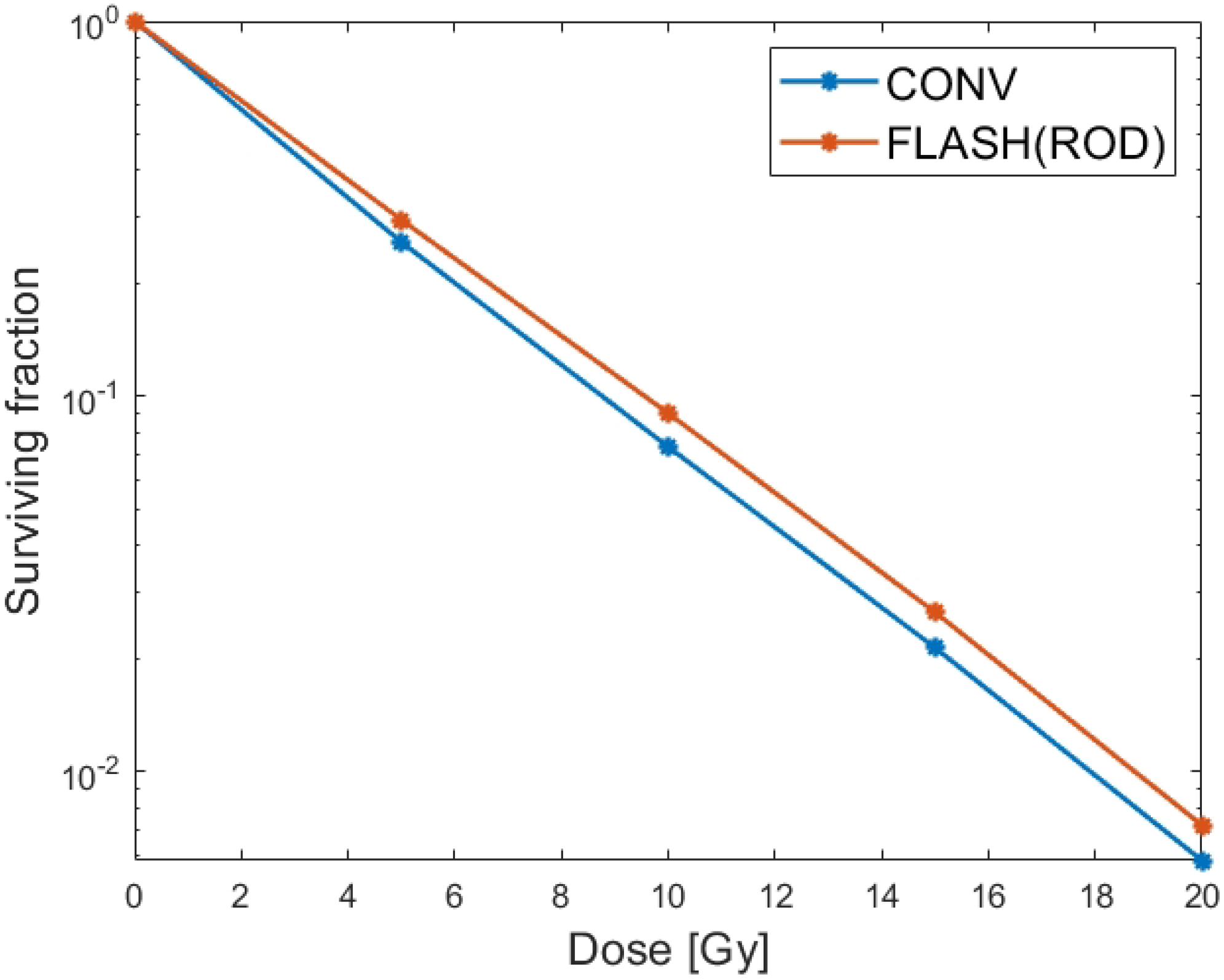
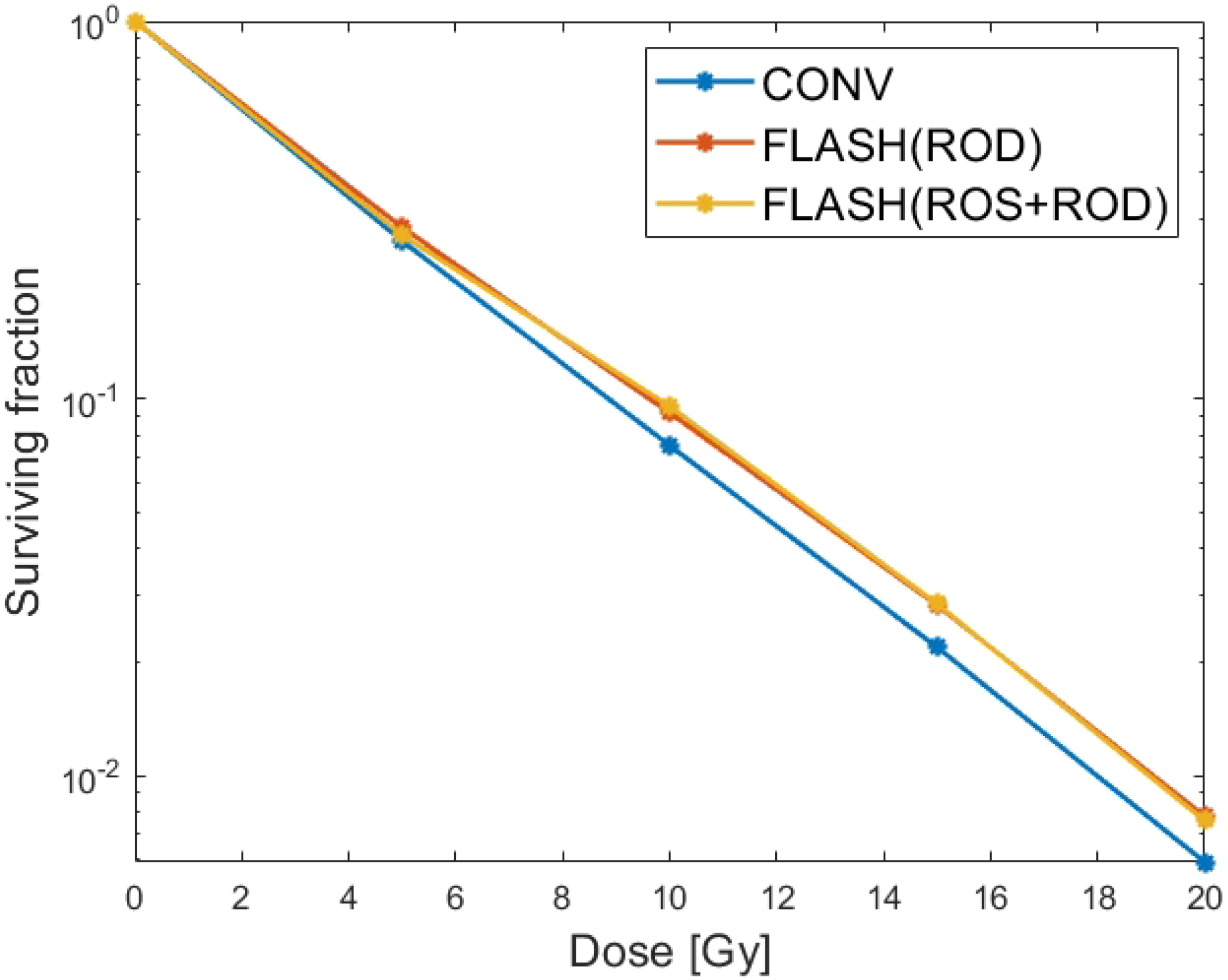
CONV-RT *v.s*. FLASH-RT with ROD only and ‘ROS+ROD’. The overall cell surviving fraction (in logarithm with base 10) with respect to different dose, i.e., 5, 10, 15 and 20Gy respectively. In ROS+ROD FLASH model, by using *S*_*ROD*_ = 15.3 *mm Hg*/*s, a* = 8, *b* = 200, *k*_*ROS*_ = 0.5 · 10^―2^ *s*^―1^ and *c*_*p*_ = 4.5 · 10^―4^ *s*^―1^, one can get similar surviving fraction as the ROD FLASH model.

As we known, Flash-RT can effectively protect normal tissues while maintaining tumor killing[34]. In order to show the differential response of FLASH-RT to tumor and healthy cells, we considered the 0-0.5mm cell model as tumor and healthy cells respectively and plotted the curves of their oxygen tension and cell survival fraction using Eq. (1), (3) and (9) in Figure 5. Compared to healthy cells, tumor cells have a higher cell proliferation rate, which also leads to higher oxygen consumption in tumor cells[35]. According to [8], in the broader range of values for *S*_*m*_ (i.e., *S*_*m*_ = 2-7 mm Hg/s), we achieved a reasonable fit for *S*_*m*_ = 3.2 mm Hg/s in healthy cells. As can be seen from Figure 5, for healthy cells, the intracellular oxygen tension is higher, and FLASH-RT has a significant protective effect on healthy cells, while for tumor cells, the intracellular oxygen tension is lower, and there is no significant difference in the surviving fraction between FLASH-RT and CONV-RT.

**Figure 5.**
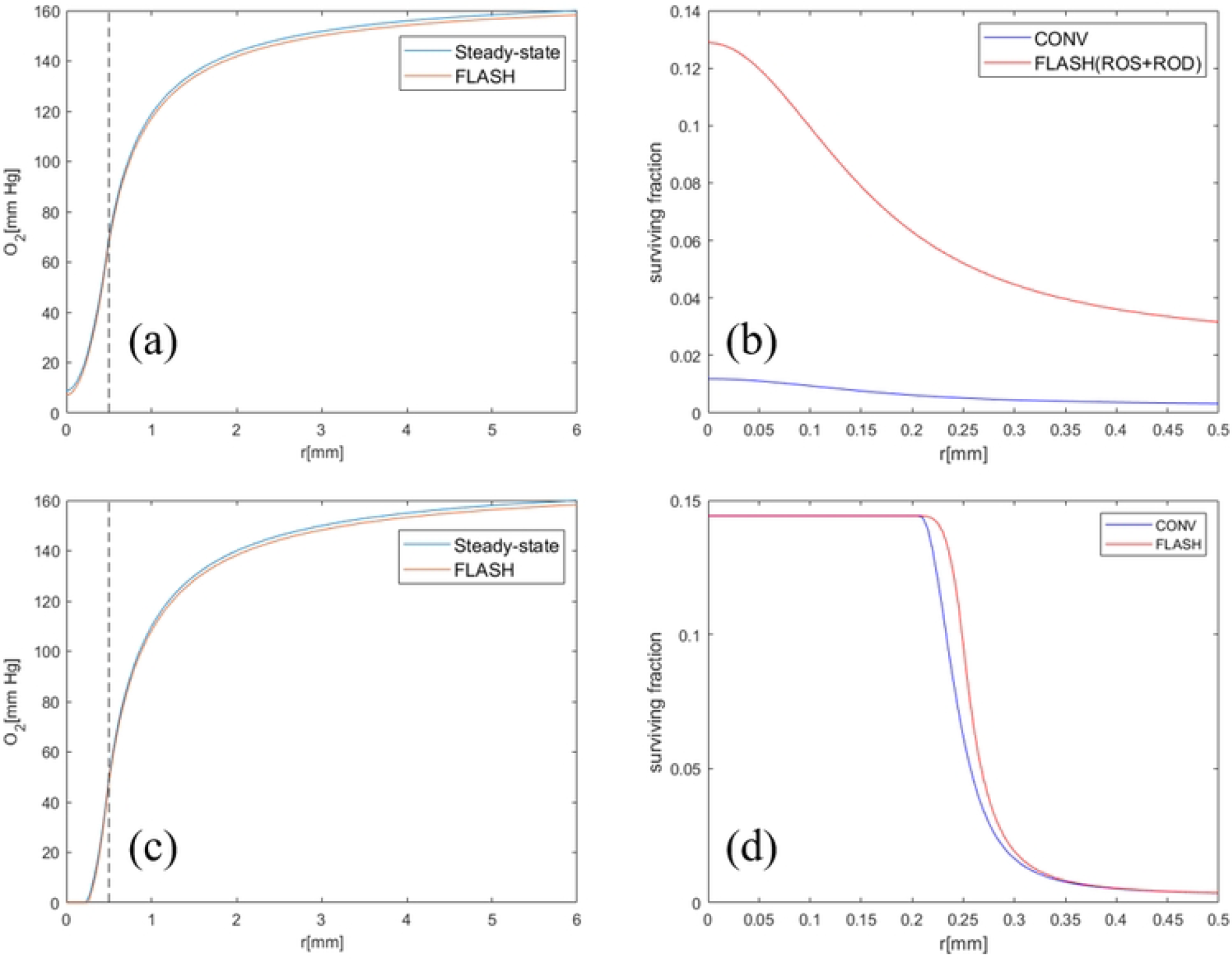
Comparison of oxygen tension and cell surviving fraction between Healthy cells and tumor at a dose of 10 Gy. (a)(b) Oxygen tension and survival fraction when all cells in spheroid model are healthy cells with R=0.5mm. The simulation parameters can refer to Table 1 and *S*_*m*_=3.2mm Hg/s and *S*_*ROD*_ = 15.3mm Hg/s. (c)(d) Oxygen tension and survival fraction when all cells in spheroid model are tumor cells with R=0.5mm. The simulation parameters can refer to Table 1 and *S*_*m*_=4.2mm Hg/s and *S*_*ROD*_ = 15.3mm Hg/s.

## 4. Discussion

We have modeled the changes of oxygen tension and ROS level in cells in order to explain the differential response of tumors and normal tissues to FLASH-RT. The previous work with ROD alone [8] can only explain the radiation protection of normal tissues owing to the reduction of oxygen tension.

In this paper, ROS was considered as a possible influencing factor. By comparing the difference of ROS levels between FLASH-RT and CONV-RT, and by simulating the changes of ROS in cells and the effect of oxidative stress on cell surviving fraction, the biological effects of FLASH-RT can be better explained. In addition, although ROS are considered in our model, some tumor cells still have high survival rate after FLASH-RT. This is because although these tumor cells will not have a protective effect from the slightly lower ROS level than CONV-RT, they still enjoy the protective effect caused by the reduction of oxygen tension. To date, the underlying mechanism for the FLASH effect has not been fully understood, so the ROS+ROD FLASH model via differential equations needs to be experimentally validated. For example, changes in ROS level may not be well characterized during FLASH-RT and CONV-RT, this makes it impossible for us to accurately evaluate the cell damage caused by the oxidative stress effect of ROS. So we can only measure the effect of ROS on cell surviving fraction by comparing the difference of ROS between FLASH-RT and CONV-RT.

A potential future work is to incorporate this ROS+ROD FLASH model into the FLASH treatment planning. That is, besides routine planning objectives, one also needs to optimize the spatiotemporal dose and dose rate distribution in order to maximize the surviving fraction for normal tissues and minimize the surviving fraction for tumors as quantified by Eq. (9), which can be more quantitative and accurate than current FLASH treatment planning methods that optimize FLASH-RT dose rate [30,31] or FLASH effective dose [32] without using mathematical models of the FLASH mechanism.

## 5. Conclusion

A new FLASH model that also accounts for ROS in addition to ROD is developed that can explain the differential response of FLASH to tumors and normal tissues.

## Data Availability

All relevant data are within the manuscript and its Supporting Information files.

## Acknowledgments

The authors are very thankful to the valuable comments from anonymous reviewers.

